# Toxic or not toxic? Interlaboratory comparison reveals almost fifty-fifty chance in the cytotoxicity assessment (ISO 10993-5) of an identical medical device

**DOI:** 10.1101/2023.03.28.23287847

**Authors:** Sarah Gruber, Angela Nickel

## Abstract

**Background:** Medical device manufacturers are obliged to prove the biocompatibility of their products when they come into contact with the human body. The requirements for the biological evaluation of medical devices are specified by the international standard series ISO 10993. Part five of this series describes the performance of *in vitro* cytotoxicity tests. This test evaluates the effects of medical device use on cell health. The existence of the specific standard suggests that the tests will produce reliable and comparable results. However, the ISO 10993-5 offers wide latitude in the test specifications. In the past, we noticed inconsistencies of the results from different laboratories.

**Objective:** To determine if the specifications of the standard ISO 10993-5 are sufficient for assessing the comparability of test results and, if not, identify potential influencing factors.

**Methods:** An interlaboratory comparison was conducted for the *in vitro* cytotoxicity test according to ISO 10993-5. Fifty-two international laboratories evaluated the cytotoxicity for two unknown samples. One was polyethylene (PE) tubing, which is expected to be non-cytotoxic and the other was polyvinyl chloride (PVC) tubing, for which a cytotoxic potential was presumed. All laboratories were asked to perform an elution test with predefined extraction specifications. The other test parameters were freely chosen by the laboratories according to the guidelines set by the standard.

**Results:** To our surprise only 58 percent of the participating laboratories identified the cytotoxic potential of both materials as expected. Particularly for PVC a considerable variation of the results between the laboratories was observed (mean = 43 ± 30 (SD), min = 0, max = 100). We showed that ten percent serum supplementation to the extraction medium, as well as longer incubation of the cells with the extract, greatly increased the test sensitivity for PVC.

**Conclusion:** The results clearly show that the specifications set by the ISO 10993-5 are not sufficient to obtain comparable results for an identical medical device. To set requirements that ensure reliable cytotoxicity assessments, further research will be necessary to identify the best test conditions for specific materials and/or devices and the standard needs to be revised accordingly.

## 1 Introduction

Medical devices have numerous applications, e.g. from dressing materials to surgical instruments to implants. To fulfill their intended purposes, medical devices are composed of various materials. Since several of them come into contact with the human body, medical device manufacturers have to ensure that their products and the materials used are safe. This is especially critical when the medical devices have long-term contact with human tissue, like implants or prostheses. Materials including additives or residues from manufacturing and/or cleaning processes may cause adverse effects such as cellular damage and allergic or local tissue reactions.

To gain market access, biocompatibility has to be proven for products that come in direct or indirect (fluid/gas transmitted) contact with the human body of either the patient or the user (1,2). The requirements for assessing biocompatibility for medical devices are specified in the international standard series ISO 10993. Depending on the specific device, as well as the intended use, different tests are necessary to confirm biological safety. Usually, *in vitro* cytotoxicity testing is required to evaluate the impact on cellular health (3). For this, either the product or more common extracts from it (figure 1) are tested in mammalian cell culture by assessing the impact on cell vitality via the analysis of cell growth, replication, and/or morphology (4,5).

**Figure 1.**
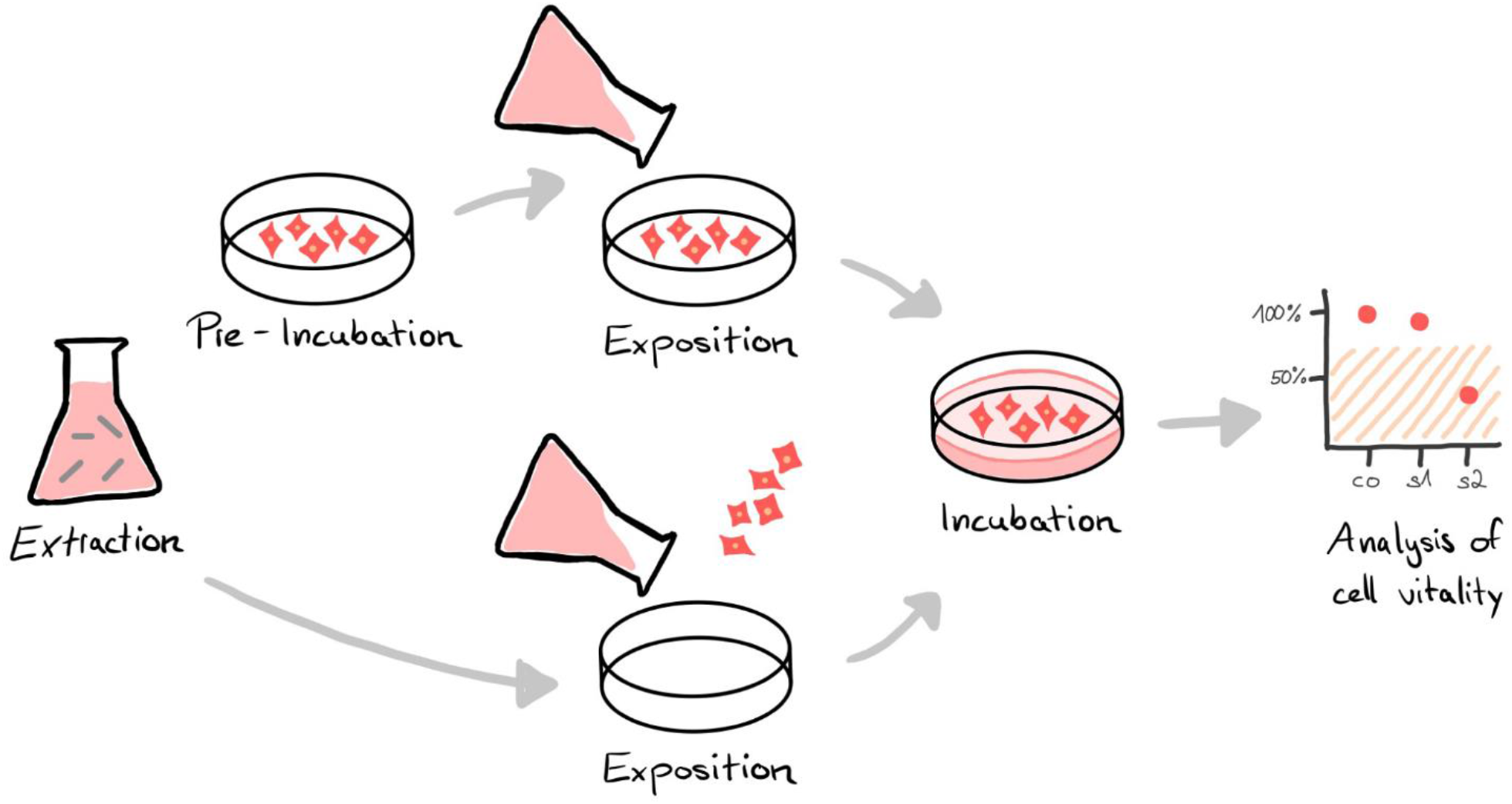
*In vitro* cytotoxicity analysis of medical devices is often performed using an elution test. Instead of testing the cytotoxic potential of the final device directly, the materials or the product to be tested are immersed in an extraction medium (extraction). The resulting extract is either exposed to a pre-incubated cell layer or cells are seeded directly in the extract (exposition). After an incubation period, the vitality of the cells is analyzed by special assays, e.g. via microscopic and/or colorimetrical evaluation (analysis). A reduction in cell vitality of the sample (s) compared to an untreated control (co) by more than 30 % is considered cytotoxic.

The specifications for *in vitro* cytotoxicity tests are described in the standard ISO 10993-5 (5). To provide sufficient coverage for various medical devices and application scenarios, the standard is intentionally kept open in many aspects (4,5). This allows test designs to be freely adapted to the clinical use scenario, such as exaggerate extraction for critical applications or different materials, as well as the nature of the medical device, e.g. substance-based medical devices. In addition, the flexible specifications facilitate adaptation to technological developments and allow adjustments to laboratory specific routines. Figure 2 provides an overview of the different parameters for *in vitro* cytotoxicity tests. Since the parameters can be combined in multiple ways, this results in a huge variety of test designs.

**Figure 2.**
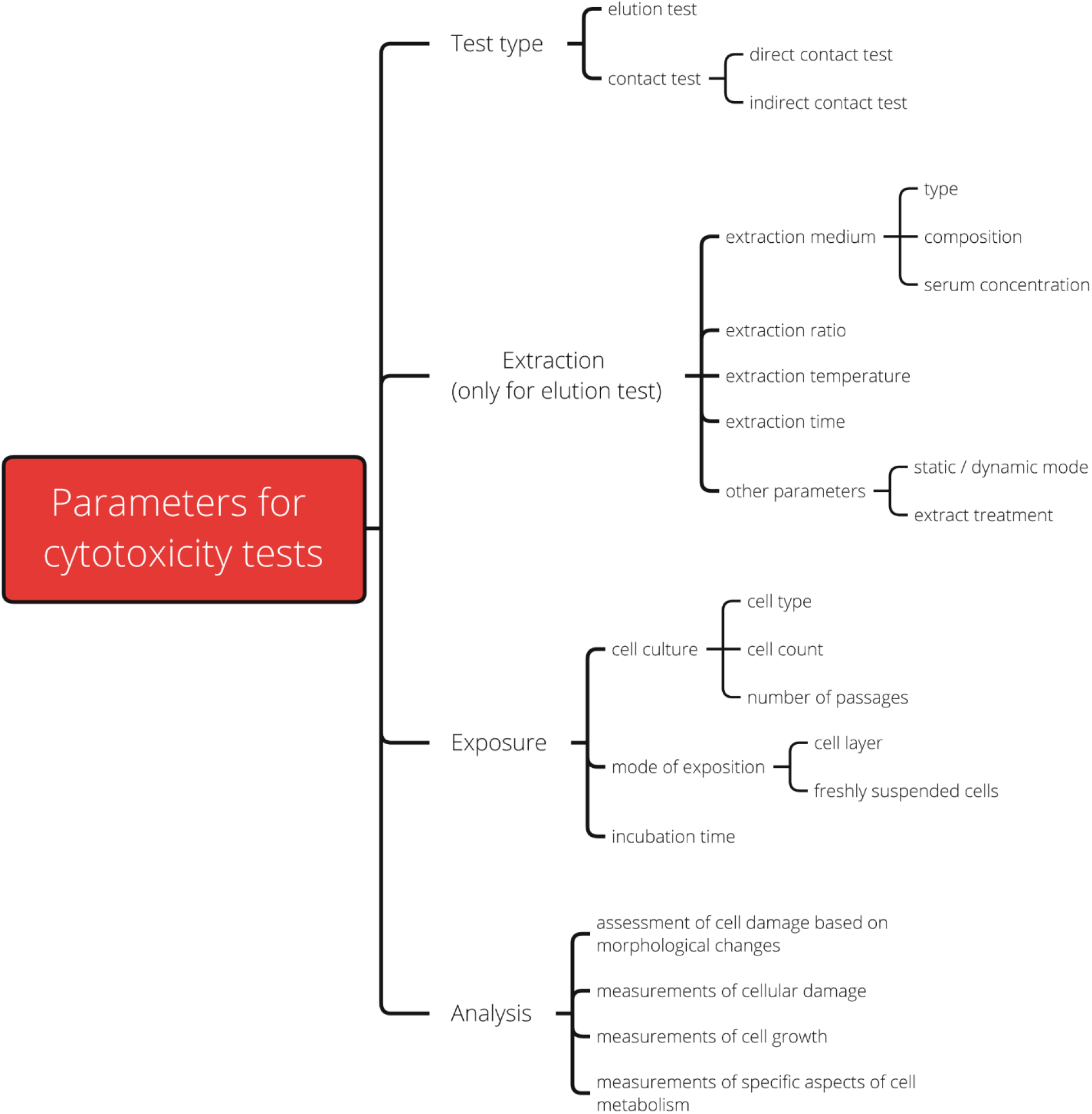
Overview of the parameters for *in vitro* cytotoxicity tests according to ISO 10993-5.

Even though the openness of the specifications is reasonable, it leads to limited comparability and unreliable test results in practice. In our work as medical device consultants, we observed that different laboratories do not always deliver identical cytotoxicity classifications for the same sample. For example, manufactures frequently receive passed cytotoxicity test certificates from the component suppliers. However, when the final product is tested again for cytotoxicity, marked cell inhibition is detected, which often cannot be explained by the assembly process. In addition, this impression is reinforced by the fact that several studies identified parameters that are particularly well suited for certain materials to determine their cytotoxicity (6–9).

To investigate these observations more closely, we conducted an interlaboratory study comparing the test results of 52 international laboratories for two provided materials: a non-cytotoxic polyethylene (PE) and a cytotoxic polyvinyl chloride (PVC). Our goal was to determine if the specifications set by the standard ISO 10993-5 are sufficient to yield comparable test results and, if not, identify potential influencing factors.

## 2 Methods

### 2.1 Interlaboratory comparison

#### 2.1.1 Scope of the interlaboratory comparison

An interlaboratory comparison was performed for the *in vitro* cytotoxicity test of medical devices to investigate whether the specifications set by the standard ISO 10993-5 are sufficient to obtain reliable and comparable results among different laboratories for the same sample. Cytotoxicity is defined as a reduction in cell viability by greater than 30 percent compared with an untreated control (5).

#### 2.1.2 Selection of laboratories

The interlaboratory comparison was conducted in two parts. First, a German national preliminary study was conducted in 2020, followed by a larger-scale international interlaboratory study in 2021. Both results were evaluated together in this report. In total, two hundred and fifty laboratories were invited globally to take part in the interlaboratory comparison. All these laboratories offer *in vitro* cytotoxicity testing for medical devices in accordance with ISO 10993-5. Fifty-six laboratories enrolled in this study. As a prerequisite for participation, the anonymity of the laboratories, as well as their test results, had to be guaranteed. The participation was voluntary and not compensated. Four of the participating laboratories had to be rated as “failed”, because they did not submit their test results. Forty-seven of the 52 successful participating laboratories were either accredited according to ISO/IEC 17025 (General requirements for the competence of testing and calibration laboratories; (10)) and/or certified according to the Good Laboratory Practice (GLP). The laboratories included in the comparison were located in 18 different countries: Argentina, Austria, Belgium, Brazil, Canada, China, Czech Republic, France, Germany, India, Italy, Norway, Poland, Portugal, Spain, Switzerland, Turkey and United States of America.

#### 2.1.3 Selection and preparation of test materials

Two testing materials with generally known cytotoxic potentials were selected (5,11):

a. **polyethylene (PE) tubing** (Bürkle™ 8878-0406), which can generally be assumed to be non-cytotoxic (reduction of cell viability ≤ 30 %) when properly manufactured and without relevant toxic residues.
b. **polyvinyl chloride (PVC) tubing** (Thermo Scientific, Nalgene® 8000-0020), depending on the formulation and embedded plasticizer, a cytotoxic potential is expected (reduction of cell viability > 30 %).

Both materials were selected for their high material quality (e.g. food grade), aiming to eliminate natural variations in the material as much as possible. Test samples were cut and sterilized with ethylene oxide (EO). To further minimize variation effects, several samples were pooled (four pieces of tubing as one sample per material). In total each final pooled test sample was 50 cm^2^.

#### 2.1.4 Pre-testing of materials

To exclude the influence of possible variations in the material, the test materials were pre-tested for homogeneity. Before the interlaboratory comparison, one GLP certified laboratory tested each of the materials for cytotoxicity ten times on different test days. The same test conditions that applied to the study participants were complied with (see 2.1.5). To ensure that variations in the applied cell viability assay setup did not influence the final assessment, several cytotoxicity measurements were obtained for each test run. No significant material variations were detected. PE was identified as non-cytotoxic (mean reduction of cell viability = 11.5 % ± 9.1 (SD)) and PVC was identified as cytotoxic (mean reduction of cell viability = 88.9 % ± 5.5 (SD); figure 3).

**Figure 3.**
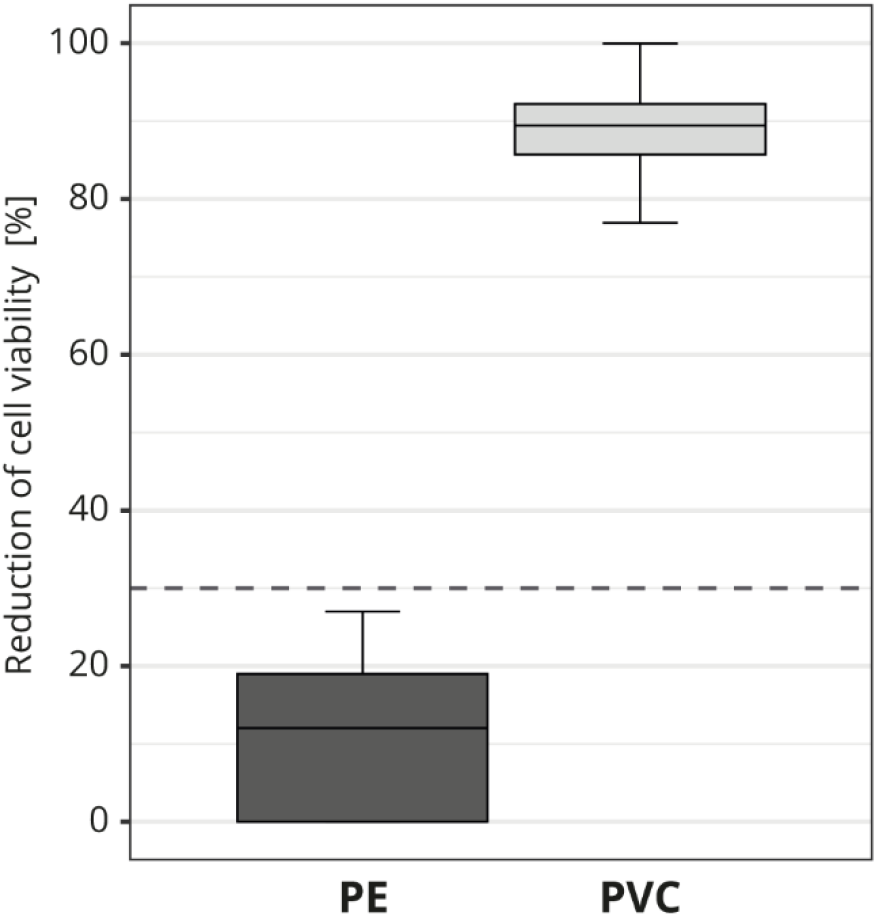
Pre-testing of the cytotoxic potential of the test materials used in the interlaboratory study. The box plot shows the measured cytotoxicity values for polyethylene (PE) and polyvinyl chloride (PVC). The threshold for cytotoxicity, a reduction of cell viability by more than 30 % compared to an untreated control, is indicated by the dotted line.

#### 2.1.5 Test procedure

Each laboratory was provided with one test sample each of PE and PVC. The specific material selection and the cytotoxic potential were not known to the laboratories. The standard ISO 10993-5 sets the requirements for *in vitro* cytotoxicity tests for medical devices. To ensure better interlaboratory comparability, some test parameters were predetermined. The laboratories were asked to perform an elution test since this procedure is the most frequently used. The contact duration of the medical device with the human body is usually simulated by the extraction parameters. Thus, to reduce variations, the main specifications for the extraction were predefined as follows: extraction ratio: 6 cm^2^/ml (surface/volume); extraction temperature: 37 °C; and extraction time: 24 hours. No further parameters were specified to allow the observation of where and how variations between laboratories would turn out. Overall, laboratories were instructed to choose the most sensitive test setup according to the guidelines set by the ISO 10993-5. The primary objective of the interlaboratory comparison was to compare the laboratories and not determine their reliabilities. Therefore, only a single test run was performed. We assumed that the laboratories had already confirmed the repeatability of their test systems.

### 2.2 Data analysis

#### 2.2.1 Data confidentiality

We intended to present the influence of as many test parameters (summarized in figure 2) as possible. However, to closely simulate real life conditions, the laboratories were asked to present their results in the form of their usual test reports. As a consequence, not all test parameters were described in full by the laboratories and made available for further evaluation. To increase the validity of the results and ensure that none of the participants could be identified, the published information density had to be partially restricted. For this reason, no fewer than three laboratories were grouped in each of the assessed categories, and the exact measured values were intentionally not shown in a readable form.

#### 2.2.2 Data analysis

Due to different test settings and different evaluation methods, the cytotoxicity values of < 0 % and > 100 % have been submitted. For better comparison, all results were set to 0 % for values < 0 % and 100 % for results > 100 %. Furthermore, the grade-based qualitative evaluations were converted to the corresponding cytotoxicity in percentage based on the grade classification; Grade 0 ≜ 0 %, grade 1 ≜ 10 %, grade 2 ≜ 30 %, grade 3 ≜ 60 %, and grade 4 ≜ 85 %.

Contrary to our study requirements, two of the successful participating laboratories did not perform an extraction but tested the materials directly. Their results were nevertheless considered in the interlaboratory comparison (with the exception of the evaluation of the extraction parameters).

A descriptive data analysis was conducted. Categorical variables were reported as relative frequencies. All box plot diagrams show the distribution of the measured cytotoxicity values. If cell viability was reduced by more than 30 % relative to that of the non-treated cells, the tested material was considered cytotoxic (5).

## 3 Results

### 3.1 Outcome of the interlaboratory comparison: highly variable cytotoxicity test results for PVC

For the interlaboratory comparison, the cytotoxic potentials of the two samples (PE and PVC) were analyzed by 52 laboratories. Figure 4 shows all individual test results for PE (A^1^) and PVC (B^1^) in ascending order. The majority (92 %; 48 out of 52) of the laboratories identified PE as non-cytotoxic (figure 4A^2^); however, only 62 % (32 out of 52) determined PVC to be cytotoxic (figure 4B^2^). Additionally, most of the values were clearly below the 30 % threshold for PE (figure 4A^1^), while the results for PVC were less distinct. The measured cytotoxicity values for PVC are much more variable and almost all possible values are represented (figure 4B^1^).

**Figure 4.**
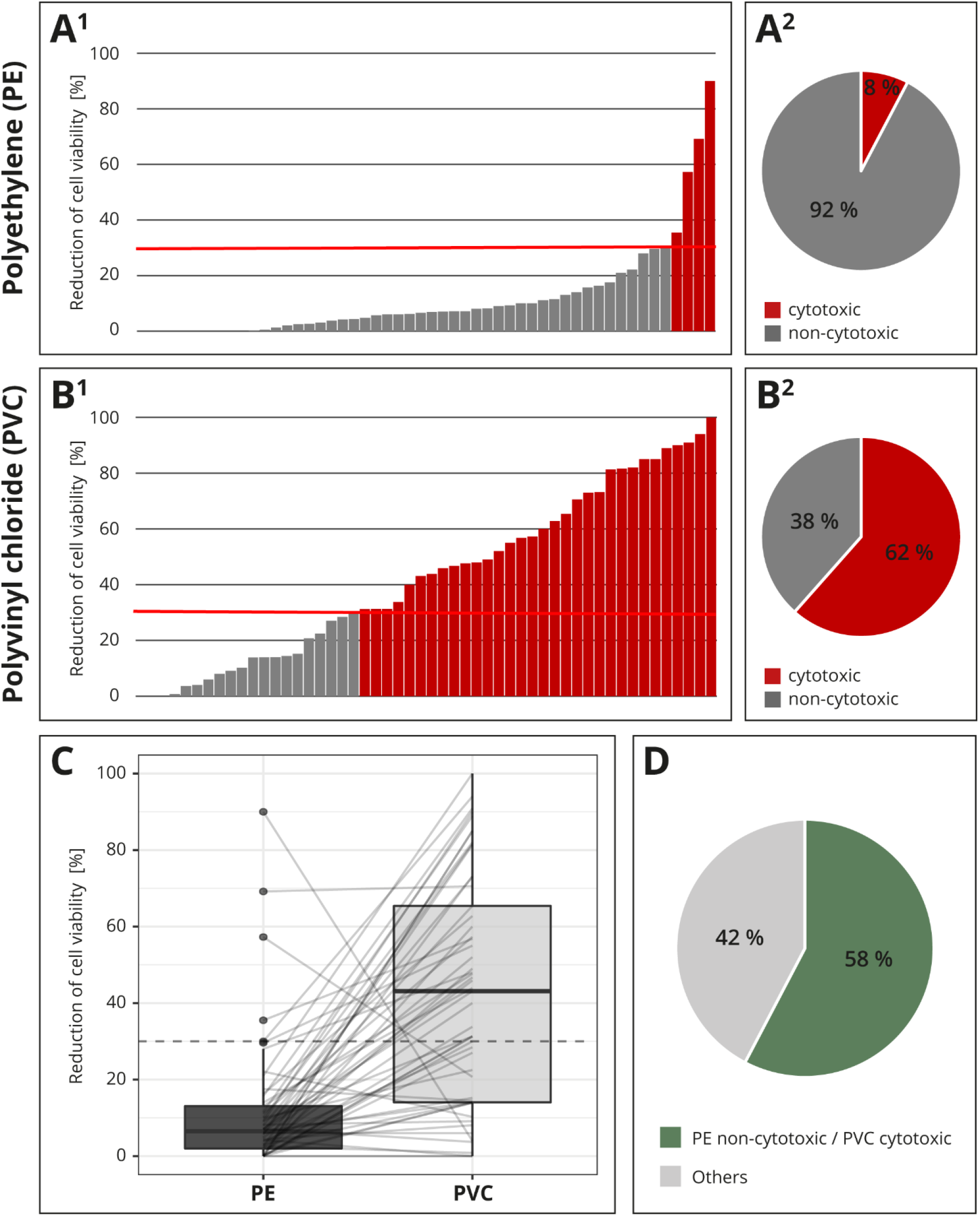
Result of the interlaboratory comparison. The threshold for cytotoxicity, a reduction of cell viability by more than 30 % compared to an untreated control, is indicated by a red or dotted line, respectively. The individual cytotoxicity test results for the materials polyethylene (PE, **A**^**1**^) and polyvinyl chloride (PVC, **B**^**1**^) were presented in ascending order and summarized in a pie chart (**A**^**2**^, **B**^**2**^). All test results were presented in a box plot and related values from each laboratory were linked with a line (**C**). Summary of whether the cytotoxic potential of both materials has been identified as expected (**D**).

For better comparison, all the results were plotted and the related values from each laboratory were linked with a line. If all samples were classified as expected, all lines would run from bottom left to top right. A horizontal line indicated a low-test sensitivity or other flaws within the setup. Several horizontal lines and even two lines running from top left to bottom right were detected (figure 4C).

Altogether, only 58 % (30 out of 52) of the participating laboratories identified the cytotoxic potentials of both PE and PVC, as expected; in 42 % (22 out of 52) of the reports, at least one of the materials was not characterized as expected (figure 4D). The accreditation or certification of the laboratories were not decisive. The distribution of results “cytotoxic” and “non-cytotoxic” for the PVC tubing among the few non-accredited laboratories was comparable to that of the accredited or certified participants (data not shown).

To investigate the factors in the test setup with the greatest influence on the observed result variations, the impact of specific parameters was investigated. For higher explanatory power and to prevent conclusions about the performance of individual laboratories, at least three test results were grouped together. The influence of specifications with less than three participants were therefore not evaluated.

### 3.2 Relevance of extraction parameters for test sensitivity

The extraction usually simulated the contact duration of the medical device. Most extraction parameters (ratio, temperature and time) were prespecified in the interlaboratory comparison. The determination of the remaining parameters, like the choice of the extraction medium was left to the participating laboratories. Only the results of the laboratories that performed an elution test (n = 50) could be included in this analysis.

Almost all laboratories (46 out of 50) in the interlaboratory comparison chose cell culture media for the extraction. Serum was often supplemented to the cell culture medium at a concentration of four to ten percent. Laboratories that did not report the serum concentration (16 out of 46) were excluded from the evaluation to avoid skewing the results. The most sensitive results were obtained with 10 % serum concentration. Only 16 % (4 out of 25) of the laboratories that added 10 % serum to their extraction medium classified PVC falsely as non-cytotoxic. However, all the laboratories (5 out of 5) that worked with less than 10 % or no serum misidentified the cytotoxic potential of PVC (figure 5A). Different serum types were not declared and could not be further analyzed.

**Figure 5.**
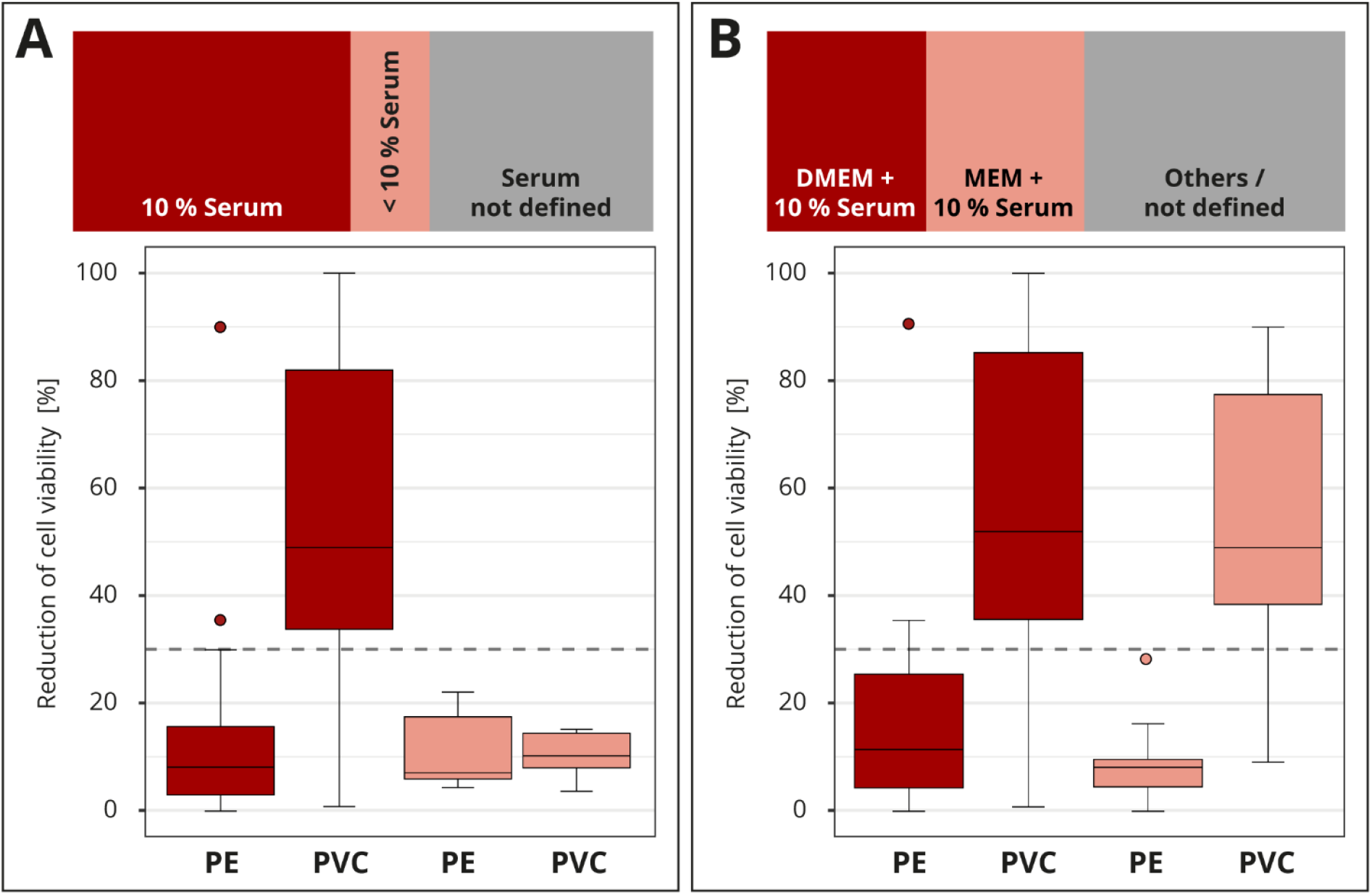
Influence of the extraction. The colors of the cuboids and the box plots represent the evaluated category. The quantitative distribution of each category is represented by the cuboids. The box plots show the measured cytotoxicity values for polyethylene (PE) and polyvinyl chloride (PVC). The threshold for cytotoxicity, a reduction of cell viability by more than 30 % compared to an untreated control, is indicated by the dotted line. Comparison of 10 % serum supplementation to the extraction medium to concentrations below 10 % in the assessment of the cytotoxic potential of PE and PVC. Data sets with unspecified serum concentration were not further analyzed (**A**). Evaluation of the cytotoxicity results if extraction media DMEM (Dulbecco’s Modified Eagle Medium) or MEM (Minimum Essential Media) plus 10 % serum supplementation were used. All other media compositions were not further analyzed (**B**).

The media DMEM (Dulbecco’s Modified Eagle Medium) and MEM (Minimum Essential Media) were the most common (39 out of 50). The rest were grouped as “others / not defined” and not further analyzed (figure 5B). Even though the compositions of the media differed slightly, no impact on the results was established. Overall, good results were obtained for all media, with a few false negative and positive results (data not shown). No differences in the cytotoxicity assessment for PE and PVC were observed when the results for DMEM or MEM plus 10 % medium supplementation were compared (figure 5B). For other extraction factors, such as whether a shaker was used (dynamic mode) or the final extract was filtered, no influence was observed (data not shown). However, too little information was provided for these parameters to allow a good conclusion.

### 3.3 Influence of cell culture setup on test sensitivity

The requirements for cell culture execution were not prespecified in the interlaboratory comparison. The majority of the participating laboratories used a comparable cell culture setup. Most laboratories (41 out of 52) worked with the mouse fibroblast cell line L929, which is one of the recommended cell lines by the ISO 10993-5. Good results were also obtained with other cell lines; however, the L929 cell line tended to produce slightly more distinct results (figure 6A).

**Figure 6.**
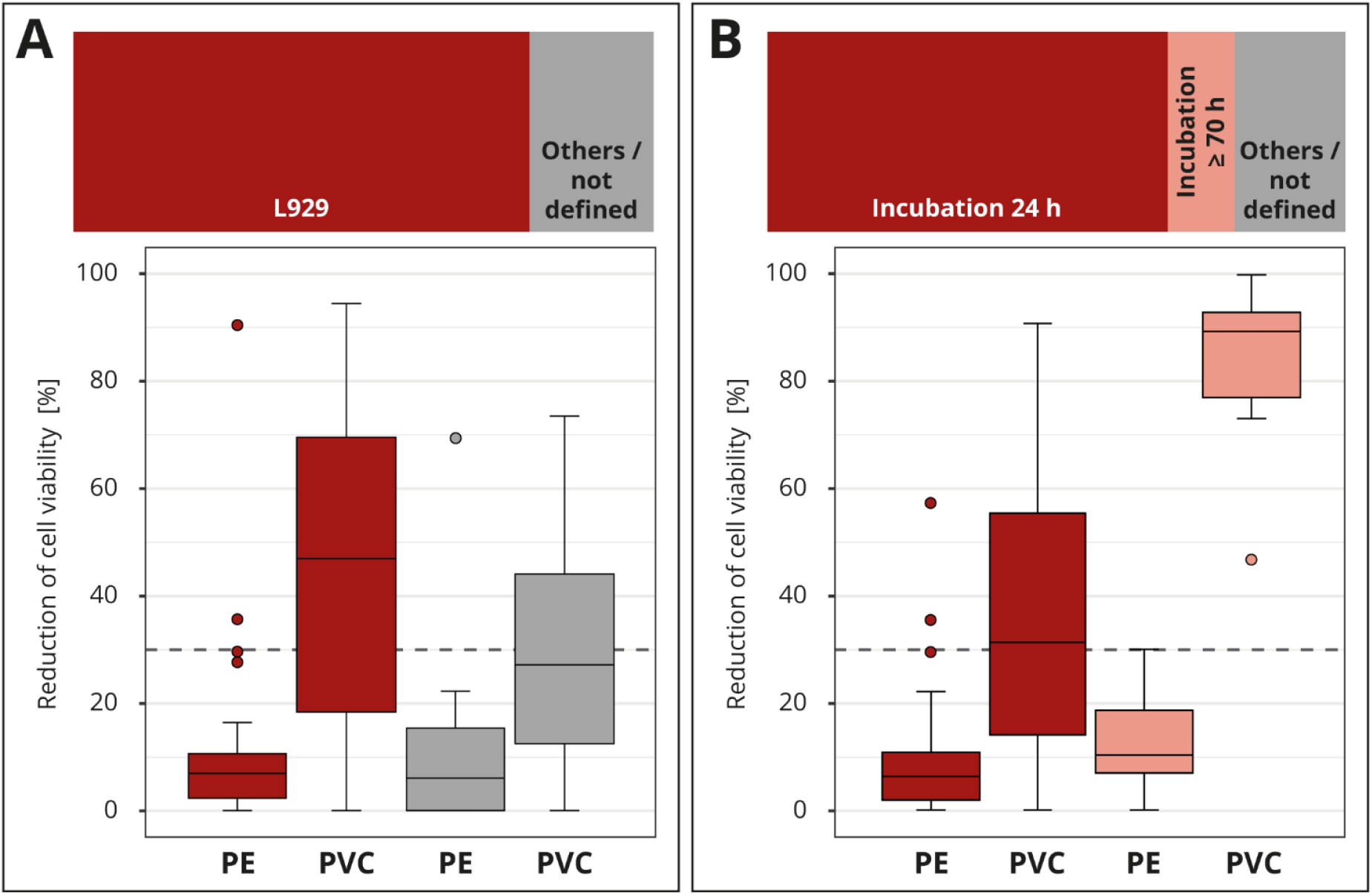
Influence of cell culture setup. The colors of the cuboids and the box plots represent the evaluated category. The quantitative distribution of each category is represented by the cuboids. The box plots show the measured cytotoxicity values for polyethylene (PE) and polyvinyl chloride (PVC). The threshold for cytotoxicity, a reduction of cell viability by more than 30 % compared to an untreated control, is indicated by the dotted line. Comparison of the obtained cytotoxicity results for the cell line L929 to all other cell lines grouped as “others / not defined” (**A**). Evaluation of the relevance of the incubation time comparing 24 and >70 hours (h). All other incubation intervals are grouped as “others / not defined” and not further analyzed (**B**).

The contact time of a medical device to the human body can be simulated in general by the extraction duration. However, there is discussion if the exposition could also be simulated by the incubation time of the cells with the extract. We are familiar with both lines of this argument. The standard ISO 10993-5 defines an incubation time of at least 24 hours or, if necessary, longer until the cells are subconfluent. The majority (36 out of 52) of the laboratories applied this minimum recommended time. A smaller group (6 out of 52) incubated the cells much longer, for at least 70 hours. The “others / not defined” group was not considered in more detail, since the information provided was not detailed enough (figure 6B). All laboratories that used the prolonged incubation time identified PVC as cytotoxic. Only 44 % (16 out of 36) of the laboratories who incubated for just 24 hours assessed PVC correctly. The results show that a longer incubation of the cells in the presence of the extract led to a clearly increased test sensitivity with regard to the determination of the cytotoxicity of PVC.

### 3.4 Impact of the cell viability assay on the cytotoxicity assessment

Different methods are available to determine the cell viability (see figure 2). The assays MTT and XTT (abbreviations based on the dyes 3-(4,5-dimethylthiazol-2-yl)-2,5-diphenyl-2H-tetrazolium bromide (MTT) or 2,3-bis-(2-methoxy-4-nitro-5-sulfophenyl)-2H-tetrazolium-5-carboxanilide (XTT)) or the neutral red uptake (NRU) assay were the most commonly (40 out of 52) used in the interlaboratory comparison (figure 7). In total, seven assays/stains (Bradford, Chrystal violet, MTT, NRU, Resazurin, Trypan blue and XTT) were used for quantitative evaluation, other results were evaluated qualitatively using grade classification.

**Figure 7.**
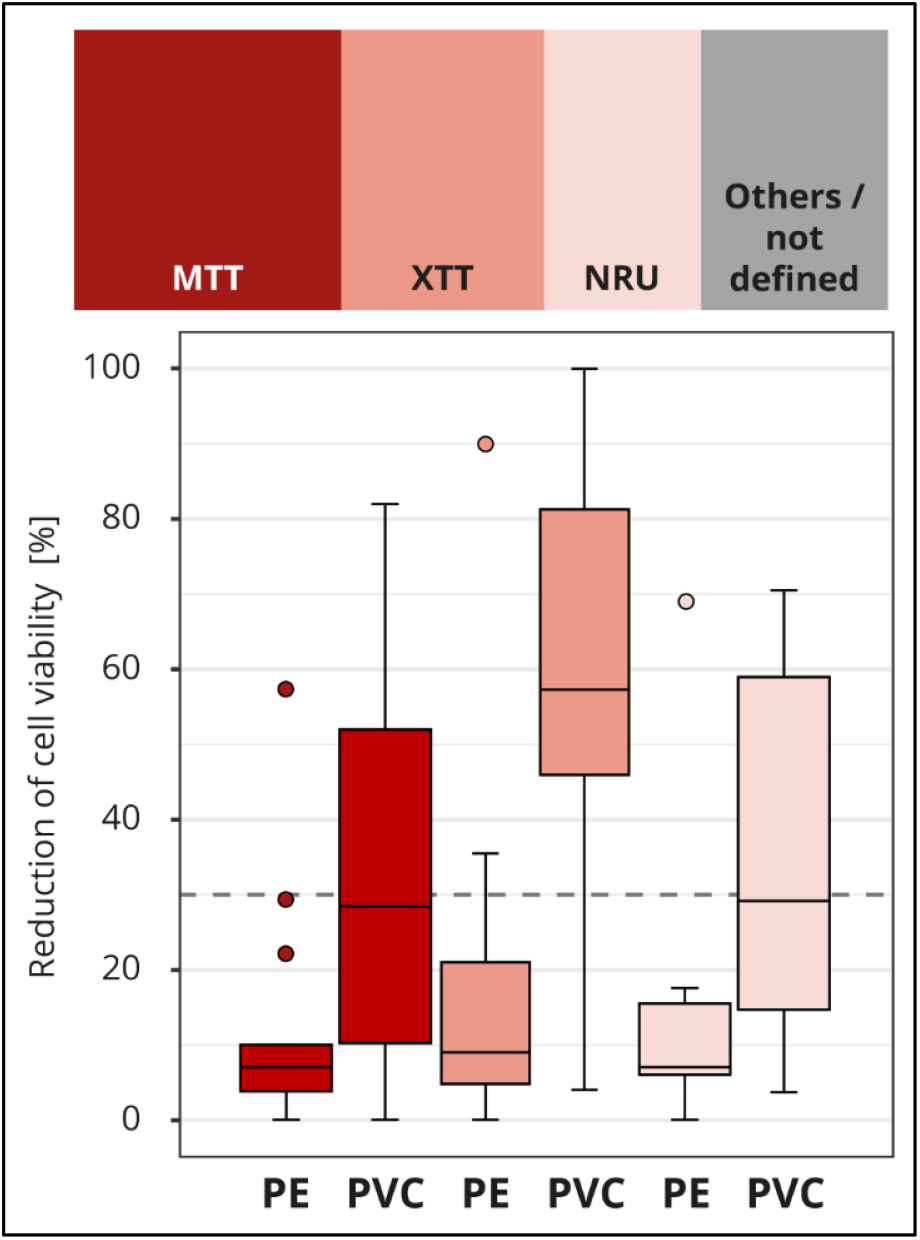
Influence of cell viability assay. The colors of the cuboids and the box plots represent the evaluated category. The quantitative distribution of each category is represented by the cuboids. The box plots show the measured cytotoxicity values for polyethylene (PE) and polyvinyl chloride (PVC). The threshold for cytotoxicity, a reduction of cell viability by more than 30 % compared to an untreated control, is indicated by the dotted line. Comparison of the obtained cytotoxicity results for the cell viability assays MTT and XTT, terms abbreviations based on the dyes 3-(4,5-dimethylthiazol-2-yl)-2,5-diphenyl-2H-tetrazolium bromide (MTT) or 2,3-bis-(2-methoxy-4-nitro-5-sulfophenyl)-2H-tetrazolium-5-carboxanilide (XTT) respectively, and neutral red uptake (NRU). All other assays/stains are grouped as “others / not defined” and not further analyzed.

At first glance, it looks like somewhat better results were obtained with the XTT assay, although just like the MTT assay, the assay mechanism depends on the cells’ metabolic activities. In contrast, no difference was found between the results of the laboratories that used the MTT or NRU assay, even though these two assays assess different aspects of cell health (figure 7). However, analysis of the test setup revealed that the differences between XTT and MTT or XTT and NRU can be explained by the laboratory’s specific choice of test parameters. In particular, the observed differences between the assays could be traced back to the amount of serum added to the extraction medium and the length of the incubation period (data not shown).

## 4 Discussion

### 4.1 Unreliable test results – a big problem for the medical device industry

Although from our experiences we expected a slight unreliability of the test results, we were surprised by the size of the problem. We did not foresee an almost 50 % chance that a tested material or device is evaluated as cytotoxic or not. PE was mostly classified, as we expected as “non-cytotoxic” and the majority of the measured values were close (see figures 5B^1^ & B^2^). Therefore, it seems that the problem rather lies with the reliable detection of cytotoxicity, in our case the correct evaluation of PVC.

PVC is very rigid and thus a brittle polymer. To make it softer and more flexible and to achieve its desired properties, plasticizers are added. These plasticizers are more or less only dissolved in the PVC polymer; thus, the molecules can leach out from the compound, which may cause undesirable consequences (12). For the PVC tested in our interlaboratory comparison, the plasticizer di(2-ethylhexyl) phthalate (DEHP) was used. Since the late 1960s numerous adverse effects have been reported, which can be associated with the release of DEHP from PVC medical devices (13,14). The use is especially critical when the exposition is either long-term or at critical points during development, such as in neonates or developing fetus (13). As a consequence, the use of DEHP, for example, has been restricted in the EU, and products containing DEHP have to be labeled (15).

Cytotoxicity has to always be addressed as an endpoint if tests are required within the scope of the biological risk assessment of a medical device, independent of the device categorization based on contact type and duration (3). Thus, it is all the more concerning if the results cannot be trusted. On the one hand, this creates uncertainty for medical device manufactures regarding the results to the safety of their products, as well as the reliability of test reports presented by suppliers. On the other hand, it is also a concern for patient safety, as unsafe products may enter the market or safe products may be kept off market, which is just as critical. In addition, several manufacturers use the cytotoxicity test as a screening test for batch releases to detect toxicologically relevant residues from production (16) to be able to respond to them quickly. If the test setup is not sensitive or reliable, problems in production may be detected (too) late. The consequences can be very serious. For example, in 2001, the company Sulzer Orthopedics recalled their hip implants after they became aware of severe complications. They traced the root of the problem to a flaw in the manufacturing process, which resulted in the incomplete removal of lubricant residue. The traces of the residue on the implant’s surface led to a loosening of the hip shell in at least 200 patients. In addition to the patients suffering, the reputation of Sulzer was severely damaged (17). Meanwhile, the current discussion in the EU due to the implementation of the new medical device regulation 2017/745 (EU-MDR (1)) illustrates the dangers that could arise from product shortages. Due to the increased requirements, niche products, such as pediatric stents or special orthopedic implants, especially, may not be available in the future, which would result in deteriorating patient care (18,19). To prevent supply shortfalls, the EU council just proposed another extension of the transition period for certain medical devices and *in vitro* diagnostic medical devices (20).

To prevent the healthcare market from being jeopardized due to unsafe or missing devices, the approval process for medical devices must be designed in such a way that sufficient safe products reach the market in a timely manner. This is why it is crucial that preclinical tests such as *in vitro* cytotoxicity testing are reliable. To ensure this, it is important that guidelines such as the standard ISO 10993-5 provide the necessary information, so that individual laboratories can set up test methods that lead to uniform results across all laboratories. However, as the results of our interlaboratory comparison showed, this is currently not the case. Hence, a deeper understanding of the factors that influence the reliability of the test results for different medical devices or materials, is needed.

### 4.2 Serum content in extraction medium affects cytotoxicity results

To identify factors responsible for the varying results in our interlaboratory comparison, we analyzed the obtained results for different parameters (extraction parameters, cell culture setup and assessment of cell viability) in more detail. For most parameters we could not make a clear statement due to insufficient data available for each characteristic. However, we clearly demonstrated the effect of serum supplementation on the intensity of measured cytotoxicity for PVC.

In 2021 Jablonská et al. (8) demonstrated for degradable metallic biomaterials how test conditions can affect the results of *in vitro* cytotoxicity testing. Among other things, they found that solutions of zinc chloride were significantly less toxic when ten percent serum was added to the extraction medium compared to five percent. This can be attributed to the serum’s protective effect since it may bind and mask toxic substances including zinc ions (5,8). In contrast, our interlaboratory comparison revealed that higher serum concentrations increased the reliability of the test results related to the cytotoxicity assessment of PVC (see figure 5A).

PVC polymers are considered inert (21), and potential cytotoxicity can be traced back to the additives. Plasticizers embedded in the PVC matrix are not water soluble. However, since serum contains various components, including proteins and lipids, it is well suited to dissolve plasticizers from the compound (22). This property may explain the higher cytotoxicity we observed for PVC, when ten percent serum was added to the extraction medium. The reason why DEHP is especially critical in medical applications, could be that DEHP migrates much faster from the PVC matrix compared to other plasticizers (12).

Nevertheless, the results from our interlaboratory comparison do not mean that PVC or even DEHP have to be considered always as concerning. Depending on the type, amount, and quality of the plasticizers added, medical devices composed of PVC can be safe for use relative to other materials. Therefore, they are rightly the gold standard in the manufacture of tubing sets for medical applications (23). Further, we should keep in mind that new materials and/or additives are not automatically superior. To ensure that improvements have been made, thorough testing is necessary, which depends on reliable test designs.

### 4.3 The standard ISO 10993-5 needs further revision

Since the publication of the standard in 1992, the ISO 10993-5 has already been revised twice, most recently in 2009 (5,24). Both revisions incorporated lessons learned from two interlaboratory studies conducted by the ISO committee. The technical report TR ISO 10993-55 describing the results from the second study conducted in 2006 with 12 participating laboratories from six countries was just released in February 2023 (24). In the latest study, all participants identified the non-cytotoxic material, a high-density polyethylene sheet, as well as two cytotoxic materials, which were segmented polyurethane films either containing 0,1 % zinc diethyldithiocarbamate (ZDEC) or 0,25 % zinc dibutyldithiocarbamate (ZDBC), as expected. ZDEC and ZDBC are recommended as reference materials for *in vitro* cytotoxicity testing (5). Even though the cytotoxic potential was identified correctly in all cases, considerable variation of the results among the laboratories were detected, especially for the cytotoxicity assessment of ZDBC (24). To reflect measurement uncertainties the threshold of at least 30 % reduction of cell viability was introduced in the standard. The authors concluded that in combination with the threshold, “the testing of the 100 % extract gives a secured finding of extractable components with a cytotoxic potential […]” (24). However, the findings from our interlaboratory comparison show that in reality this assumption is not always true. Instead, the results seem to depend strongly on the material under investigation. Therefore, we propose that further research into the interrelationships and another revision of the standard is required.

Even though we understand the reasons why the standard ISO 10993-5 is deliberately kept unspecific for several aspects and recognize the advantages (see introduction), the unreliability of the test results and, in consequence, the possible problems for patients, users, as well as medical device manufacturers are not acceptable. Nevertheless, it cannot be the responsibility of the individual manufacturer to repeatedly run the same test to optimize test settings for each individual medical device and/or material composition, especially when standard materials for medical devices are used. Rather, reliable standard test setups are needed to improve trust in the obtained results. We want to emphasize that we do not believe that the observed unreliability of the test results is principally to blame on the participating laboratories as they worked according to ISO 10993-5. Nonetheless, it is of concern that at least two laboratories probably mixed up the samples, since their cytotoxicity assessments for PE and PVC yielded opposite results to what was expected (see figure 4C). However, the main problem seems to be that the guidelines are not precise enough.

It would be ideal to have one standard *in vitro* cytotoxicity test setup that would be sensitive for all medical devices independent of the material composition as well as other treatments, such as cleaning processes or special material finishes. However, the contradictory effects we found for high serum addition compared to the research findings from Jablonská et al., 2021 (8) already indicate the unrealistic nature of this. Therefore, the goal should be to find, on the one hand, as many parameters as possible that can be used universally and on the other hand to identify factors that need to be more material specific. To do this, the following three overarching steps of the test are discussed: extraction, cell culture setup (exposition and incubation), and evaluation of cell vitality (analysis).

#### Step 1: Extraction

Extraction is one of the major influencing factors, and ISO 10993-5 and ISO 10993-12, the part for sample preparation and reference materials (25), already provide detailed information on the extraction conditions, such as the extraction duration and ratio. To prevent a major fluctuation in the results, these two parameters were specified in the interlaboratory comparison.

Another influence on extraction is the extraction medium. The ISO 10993-5 states that “the choice of the extraction vehicle(s) taking into account the chemical characteristics of the test sample shall be justified and documented.” Even though we did not provide any specifications in this regard, almost all laboratories used cell culture medium for extraction but with differences in the serum supplementation. According to ISO 10993-5, cell culture medium with serum is the preferred extraction medium because it can extract polar and nonpolar substances. However, no further information has been provided on the optimal serum concentration. As discussed, we found that ten percent serum concentration increased the test sensitivity for PVC, probably because more of the plasticizer was dissolved from the matrix. However, not all materials react the same way to serum, as shown by Jablonská et al., 2021. The serum’s protective effect, which was argued by Jablonská et al., is also mentioned in the ISO 10993-5.

Although the authors of the ISO 10993-5 acknowledge that different materials require distinct extraction conditions, no further guidance on how to determine or, at least, verify the suitability of the extraction medium for a specific material/medical device is provided. To improve the reliability of the cytotoxicity test results, further investigations analogous to the research by Jablonská et al. in 2021 and other research groups is vital to identify optimized extraction setups for standardized medical device material classes.

These findings should be included in a renewed revised version of the ISO 10993-5. Nevertheless, we should be careful about the specifications considered as optimal. The test setup that leads to the highest toxicity does not automatically need to be the best one. Instead, the setup should be as close as possible to the *in vivo* conditions the medical device will be used under, because materials/devices that are not critical can be misidentified as hazardous and, in the worst case, will not be available for patient care.

#### Step 2: Cell culture setup

Since it is the purpose of the *in vitro* cytotoxicity test to evaluate the cells’ response to the presence of toxic substances in a standardized setting, it can be assumed that the steps following the extraction do not require a material-specific setup. In principle, to avoid varying conditions for the usage of established cell lines is preferable (5). The ISO 10993-5 gives recommendations for several well-suited cell lines, such as L929 used by most laboratories in the interlaboratory comparison. For the cells to react sensitively, a high level of cell health as well as the application of good cell culture practice is a prerequisite (5). However, the standard does not sufficiently cover how different setup parameters can influence the final cytotoxicity result. For example, we showed that a prolonged incubation period in addition to ten percent serum supplementation further increased the test sensitivity for assessing the cytotoxicity of PVC. This is probably due to the fact that the cells were exposed to the toxin for a longer period.

To ensure that sensitive results can be obtained with the selected cell culture setup, validation is essential. In the informative annexes, the ISO10993-5 provides some information on validation, such as examples for positive and negative control materials. The standard states minimum upper and lower acceptance criteria; however, no definite effect levels are specified for most materials. As a result, the laboratories use positive controls; however, without corresponding reference values, this is only of limited significance. Thus, the ISO 10993-5 should specify all recommended reference materials for validation, such as the effect level, indicated by IC50 values (half maximal inhibitory concentration), that can be expected in a working test setup. In addition, the results from our interlaboratory comparison indicate that control materials are not universally suitable. Rather the controls should come from the same material family as the test substance, at least if an elution test is used, to ensure comparable toxin extraction as well as cell reaction. This correlation is already implied in the standard, although without precise specifications.

The relationships and influences discussed here should be further investigated and the results should be clearly presented in the standard. Further, it should also be reconsidered whether the informative annex is the right place for this information, as the binding nature is not clearly communicated in this way. Only more precise specifications can ensure that all laboratories validate their cell culture setup in a comparable way, thus enabling reliable test results.

#### Step 3: Analysis

To rate the toxicity of a sample, the viabilities of treated and untreated cells should be compared. Cell vitality can be assessed by the categories, cell health esp. morphology, cell growth, and cell metabolism (5). For the evaluation, the ISO 10993-5 lists several methods and assays (see figure 2), including XTT, MTT and NRU. These assays are validated standard methods that have been established over several years and are used beyond the *in vitro* cytotoxicity test for medical devices. MTT and XTT are both colorimetric assays to detect metabolic activity via a chemical color reaction that allows the assessment of the viability of the cells (26,27). With the NRU assay, cell viability is measured via the quantitative uptake of the neutral red dye into the cell, which depends on the health of the cell membrane (28). Even though the methods and assays target different aspects of cell health, such as different target proteins, we showed that an impairment of cell health is reflected in the measured values, regardless of the precise damage. Therefore, the observed unreliability cannot be attributed to the analysis and editing of the specifications is not necessary. For individual cases, however, the assay specific read-outs may have to be considered.

Even though the *in vitro* cytotoxicity test is not supposed to be a clear pass or fail test (5), in practice the statement “cytotoxic” or “not cytotoxic” according to ISO 10993-5 is usually considered sufficient, and test results are often compared without further considering the parameters. To allow for better comparability, laboratories should state the chosen parameters in more detail. The standard should also provide more precise requirements to reduce variations in the test reports.

Nevertheless, the *in vitro* cytotoxicity test is also used as a single screening or batch test; for instance, it can be used to assess the final cleaning of a product. It must detect the lowest concentrations of toxic substances and be repeatable across laboratories to prevent similar scenarios such as the hip implant scandal.

## 5 Conclusion

Patient care highly depends on the availability of good and safe medical devices. However, this goal can only be achieved if the test methods used to assess the performance and safety of the devices produce accurate and reliable results. An almost fifty-fifty chance, nearly the same probability as for a coin toss, we observed for the cytotoxicity assessment for PVC in our interlaboratory comparison cannot be acceptable. Due to its advantages (29), the *in vitro* cytotoxicity test is key in the biological evaluation of medical devices. This is especially true when it is used as we recommend as part of a step-wise approach in line with ISO 10993-1, which includes among others the assessment of data sheets and literature in combination with further analytical methods, such as chemical analyses according to ISO 10993-18.

However, the cytotoxicity test only fulfills its important role if reliable and comparable results can be ensured. Therefore, we suggest that the standard ISO 10993-5 should be revised and more research is necessary to improve the knowledge about optimized test setups for different standard medical devices and materials. Further, awareness should be increased that individual test parameters can have marked influence on the result and it is important that they are clearly stated in the test report. Without these necessary improvements, we put patients at risk for products are unknowingly unsafe or good products are falsely not gaining market access.

## Data Availability

The participation of the laboratories in the interlaboratory comparison depended on the guarantee that their identity would not be disclosed and that no statements are made about individual performance. For this reason, raw data cannot be shared and test results were aggregated in groups with at least 3 participants.

## 7 Funding

The interlaboratory comparison received no external funding. Participating laboratories were offered to contribute to the cost of the interlaboratory comparison by purchasing a more detailed report.

## 8 Acknowledgments

We would like to express our gratitude to the participants of this interlaboratory comparison and Christian Johner for supporting us. Further, we acknowledge our former colleague Hendrik Rudolph who created the basis for the international interlaboratory comparison with the Germany-wide preliminary study. We thank Heidi Seibold for assistance with some of the statistical analysis. We would also like to appreciate the efforts of our colleagues Anja Segschneider, Bettina Martin, Hanna Lutz, Manuel Baur, and Nadine Jurrmann who proofread the manuscript.

## 9 Conflict of Interest

Both authors are employed by the Johner Institute. The Johner Institute consults for medical device and IVD manufacturers as well as other stakeholders and provides training in this area. It does not develop and/or market its own medical devices. In the field of biocompatibility, manufacturers are supported from the choice of materials to the definition of test parameters and toxicological evaluation. The mission of the Johner Institute is to enable the provision of medical devices by, among other things, improving the regulatory system. To support evidence-based regulation, activities in the area of regulatory science are conducted, such as this interlaboratory comparison. The current article is not expected to result in any financial advantages.

